# Circulating inflammatory proteins predict dementia risk, are linked to structural brain changes and reveal risk mediation pathways

**DOI:** 10.1101/2025.08.29.25334731

**Authors:** Dorsa Abdolkarimi, Yue Liu, Lachlan Gilchrist, Sara Calhas, Sheena Waters, Charles R Marshall, Petroula Proitsi

## Abstract

**Introduction:** Systemic inflammation has been identified as a key factor in neurodegeneration but the value of circulating inflammatory proteins in dementia risk prediction and their causal role has not been elucidated.

**Methods:** We leveraged proteomic data from 43,685 UK Biobank participants to investigate associations between 728 Olink inflammatory proteins and incident dementia using Cox proportional-hazards (Cox-PH) models. We used Cox-PH with LASSO regularisation to calculate a sparse signature of inflammatory proteins (ProSig) predicting incident dementia. Linear regressions assessed the association between ProSig and individual proteins with brain image-derived phenotypes and Brain Age in participants with available neuroimaging data (n = 4,106). Formal mediation analyses investigated whether inflammatory proteins mediated associations between genetic and modifiable risk factors and dementia outcomes. Mendelian randomisation (MR) tested the causal relationship between inflammatory proteins and dementia outcomes.

**Results:** 218 inflammatory proteins were individually associated with incident dementia in Cox-PH models (*p_FDR_* < 0.05). A 20-protein signature significantly improved the prediction of incident dementia beyond known risk factors. TNFRSF11B, a protein linked to vascular damage, was associated with both incident dementia and reduced hippocampal volume. Two proteins, sFRP4 and MEPE, were linked to reduced Brain Age, with sFRP4 also being protective against dementia. Mediation analyses suggested that TNFRSF11B, APOE, and C7 partially mediated the effects of modifiable risk factors on dementia. MR analyses highlighted potential protective causal roles for TNFSF13 and IL17D.

**Conclusions:** By triangulating evidence, this study shows that inflammatory proteins improve dementia risk prediction and play heterogeneous causal roles in dementia pathophysiology.

## Introduction

With an ageing worldwide population, the number of people with dementia has amplified and is projected to reach 152·8 million cases by 2050 (1). Neuroinflammation is a prominent characteristic of dementia and contributes to neurodegeneration (2). Alzheimer’s disease (AD) is characterised by deposition of β-amyloid (Aβ) plaques and neurofibrillary tangles – both pathologies are exacerbated by neuroinflammation (3).

Microglia, the resident macrophages in the brain, adopt a pro-inflammatory state in response to Aβ to remove plaques; however, prolonged activation impairs their clearance capacity and contributes to neuronal damage (3). Systemic inflammation has also been linked to cognitive decline; elevated serum levels of pro-inflammatory cytokines have been associated with impairment in cognition and memory (4,5). Moreover, pro-inflammatory circulating cytokines can penetrate the blood brain barrier (BBB) either directly via transport proteins or indirectly through stimulating circumventricular organs to release pro-inflammatory cytokines that can diffuse into the brain (5,6). Alternatively, serum cytokines can stimulate afferent nerves to propagate the inflammatory responses to the brain (7). Once the systemic pro-inflammatory responses reach the brain, they can activate microglia and astrocytes to also produce pro-inflammatory mediators (5). Evidence from genome-wide association studies (GWAS) of AD and related dementias also demonstrated strong implications of immune-related genes, particularly TNF-mediated signalling pathways, in disease pathophysiology (8). Furthermore, several epidemiological studies suggested the use of non-steroidal anti-inflammatory drugs was associated with a lower risk for AD (9).

Given the decades-long trajectory of dementia, there has been a focus on identifying protein biomarkers in middle aged adults which can hint at dysregulated pathways before the presentation of disease manifestations. Recent studies have been exploring proteomic signatures associated with the incidence of dementia (10–12). However, despite the relationship between systemic inflammation and the consequent neuroinflammation and cognitive decline, the link between circulating inflammatory proteins and dementia risk remains largely understudied.

To address this gap, we have utilised the large-scale proteomic data from the United Kingdom BioBank (UKB) prospective study to examine the individual and collective associations between circulating inflammatory proteins and incident dementia. We firstly, created a sparse proteomic signature (ProSig) of inflammatory proteins predicting incident dementia. We then investigated links between this signature and individual ProSig proteins with brain image-derived phenotypes (IDPs) and Brain Age to uncover early molecular and physiological pathways that precede dementia diagnosis. We explored whether ProSig or its proteins mediate the effects of genetic (*APOE-ε4* genotype and polygenic risk (PRS) for AD and modifiable risk factors (for example smoking status and diabetes) with dementia. Finally, we investigated whether any of the ProSig proteins are causally linked with dementia. By using a triangulation framework—incorporating observational analyses, machine learning, and Mendelian randomisation—we aimed to systematically characterise the contribution of systemic inflammation to dementia risk and to identify key circulating inflammatory proteins that may serve as early biomarkers, mediators of known risk factors, or potential causal drivers of dementia.

## Methods

### Study population

The UKB is a large population-based prospective study that recruited over 500,000 participants aged 40-69 years between 2006-2010 across the UK at 22 assessment centres (13). At baseline visits, participants completed questionnaires on their lifestyle and socioeconomic background. Physical measurements, as well as blood, urine and saliva samples were collected. The UKB study leverages linked healthcare records, follow-up questionnaires, biomarker identifications, genome-wide sequencing, and genotyping to elucidate the determinants of diseases, thereby providing invaluable insights into disease aetiology and risk factors. This research was conducted using the UKB Resource under Application Number 78867.

### Proteomics

The plasma proteomics of ∼55,000 UKB participants from baseline blood samples has been characterised by the UKB Pharma Proteomics Project (UKB-PPP) (14). The study utilised the Olink Explore 3072 technology, which is an antibody-based proximity extension assay involving pairs of antibodies binding specific protein targets. The high-throughput technology allows for the simultaneous measurement of up to 3072 protein biomarkers in a single sample (14). Protein expression was qualified on a relative scale, where values were normalised, log2-transformed and reported as arbitrary Normalised Protein Expression (NPX) units (14). To date, there are over 2900 unique protein analytes identified through multiplex proteomics assays and are broadly categorised within the inflammatory, neurological, cardiometabolic or oncological Olink panels (14). The proteomic data used in this study is from the inflammatory Olink Panels (inflammation I and inflammation II).

Of the 2919 proteins available for baseline, we excluded those with >20% of missing values (n = 4), leaving 2915 proteins. Furthermore, we excluded individuals with >20% of proteomic data missing (n = 9,078). Missing values in the remaining data (n = 44,150) were imputed using the k-nearest neighbours (k=10) algorithm (“impute” R package (Version 1.72.3)) to balance bias and variance and in line with previous UKB proteomics imputation approaches (15,16).

### Disease incident and phenotypes

#### Disease incident

Dementia diagnoses were determined based on linked hospital inpatient records, linked primary health care records, self-reports and death certificate registers. Diagnosis dates were based on the earliest record from specified data sources, and time to diagnosis was calculated in years from the baseline assessment to first recorded onset. Time to diagnosis for controls was treated as a censored event and was reported based on death or the latest follow-up (August 2024). Participants who were already diagnosed with dementia at baseline appointment (prevalent cases, n = 255) or were diagnosed with dementia before the age of 60 (early-onset, n = 49) were excluded from all analyses. Primary analyses were performed on all-cause dementia (ACD), as subtype classification is dubious in UKB and the proteomics subsample is underpowered for stratified analyses (17).

#### Neuroimaging

A total of 113 IDPs including both global (n = 12) and regional (n = 101) brain volumes were obtained from T1-weighted magnetic resonance imaging (MRI) brain scans. The images were derived from Freesurfer ASEG (categories 190 and 192) and were quality-controlled and pre-processed according to the UKB imaging pipeline (18). Brain Age is a neuroimage-derived variable that can predict cognitive decline and risk of neurodegeneration (19). Brain Age was computed using a Gaussian process regression model trained on the raw T1-weighted images collected during the first imaging session, and Brain Age delta was calculated as the difference between Brain Age and chronological age (20).

#### Covariables

We adjusted for a range of non-modifiable and modifiable risk factors in our analyses. These included sex, age, ethnicity, Townsend deprivation index, education, alcohol consumption, smoking status, body mass index (BMI), prevalent hypertension and diabetes, recorded at baseline visit. We also adjusted for the number of *APOE-ε4* alleles and the estimated globular filtration rate (eGFR) at baseline visit to normalise the excretion of proteins for each individual. Self-reported ethnicities were broadly categorized as White, Black, South Asian and others according to supraordinate UK Census categories. The Townsend deprivation index reflects on the material deprivation by integrating factors such as home-ownership, employment and household overcrowding. Education was categorised as secondary education and no secondary education. Alcohol consumption and smoking status were categorised as current, former, or never. BMI was calculated as weight (kilograms) at baseline divided by height (meters) squared. Hypertension was defined using ICD-10 codes, self-reported medication use, or a mean blood pressure of ≥140/90 mm Hg at baseline assessment (21). Diabetes at baseline was assessed through ICD-10 codes, self-reports or HbA1c concentrations ≥ 6.5% (22). The UK Biobank data were previously genotyped, and *APOE* allele variants were determined from two single nucleotide polymorphisms (rs429358 and rs7412). These SNPs were extracted from imputed BGEN data using PLINK, the haplotypes were phased, and participants were subsequently classified as ε4 heterozygotes or homozygotes (23,24). eGFR was estimated from serum creatinine using the CKD-EPI equation, which accounts for age, sex, and ethnicity (25). Individuals with missing *APOE* genotype were excluded (n = 416) and the remaining missingness in other covariates were imputed using the k-nearest neighbours (k=10) method with the “impute” R package (Version 1.72.3) (15).

### Statistical analyses

Statistical analyses were performed in R Version 4.4.1 (R Project for Statistical Computing). This research utilised Queen Mary’s Apocrita HPC facility, supported by QMUL Research-IT (26).

#### Cox proportional hazards models

The association between plasma protein levels and incident dementia was examined by Cox proportional hazards (Cox-PH) models using the “survival” R package (Version 3.5-8) (27). Incident dementia referred to dementia diagnosis after baseline visit. Participants were censored at the time of dementia onset, death, or final follow-up, whichever occurred first. Prior to analysis, protein NPX values were inverse rank normalised to have a mean of 0 and a standard deviation of 1. To establish the extent to which associations might be confounded by non-modifiable and modifiable risk factors, four covariate-adjusted models were used to assess the individual hazard ratios (HR) of each protein. The basic Cox-PH model (model 1; M1) was adjusted for sex, age, age^2^ and their interactions (age×sex, age^2^×sex) at baseline. Model 2 (M2) was additionally adjusted for *APOE-ε4* allele count. Model 3 (M3) included adjustments for sex, age, age², their interactions, modifiable dementia risk factors (education, ethnicity, BMI, diabetes, hypertension, smoking, Townsend deprivation index, and alcohol consumption), and kidney function (eGFR). Model 4 (M4) adjustments include those in M3 and *APOE-ε4* allele counts. The *p*-values from all models were adjusted for false discovery rate (FDR). Moreover, to improve comparability with other studies, we applied a secondary Bonferroni-adjusted threshold based on the number of principal components accounting for 90% of the data variance.

#### Proteomic Signature (ProSig)

To find a proteomic signature predicting incident dementia, we applied Least Absolute Shrinkage and Selection Operator (LASSO) regularisation to a Cox-PH model adjusted for proteins. LASSO uses a regularisation penalty controlled by two hyperparameters: λ, which determines the degree of shrinkage applied to the coefficients, and α, which determines the weight of the penalty term. We excluded individuals with neuroimaging data from the following analyses. For hyperparameter tuning and feature selection we used the “caret” R package (Version 6.0-94) (28) to ensure proportionate allocation of cases, and partitioned 50% of the data for training, 25% for validation and 25% for testing.

Protein levels within each subset were inverse rank normalised separately. Using the training data, we performed 10-fold cross-validation with a fixed α of 1 using the “glmnet” R package (Version 4.1-8) and identified the optimal λ value over 100 iterations (29). In each iteration, we used the model with the best λ to obtain regularised coefficients for each protein. We then took an average of the regularised coefficients across all iterations and ranked proteins based on the absolute value of their average coefficient. To optimise the number of proteins in the ProSig, we incrementally added proteins based on their ranked coefficients, starting with the top-ranked protein, then the top two, and so on – generating a ProSig for the validation data at each step. We then used these incremental ProSig as a predictor in Cox-PH models assessing time to incident dementia in the validation sample. For each model, we calculated concordance statistic (C-index) and used “ggplot” in R (Version 3.5.1) to visualise the C-indices from each model against the number of proteins used for ProSig calculation and visually inspected where the plot plateaus (30). In parallel, we compared the statistical difference in the C-indices of models generated from the top 5, 10, 20 and 25 proteins with the “compareC” R package (Version 1.3.2) which utilises closed-form variance estimator and Z-statistics (31). We selected the number of proteins based on the first instance where the addition of more proteins did not yield a significant increase in the C-index.

Finally, we used the coefficients of proteins calculated in the previous step to generate ProSig for the test data. We subsequently fitted the Cox-PH models from the previous step (M1–M4) to the test data, both with and without adjustment for ProSig. The C-index for each model, with and without ProSig, was then compared to evaluate its impact on predictive performance using “compareC” R package. We conducted sensitivity analyses by restricting dementia cases to those with onset within 5, 10, and 15 years, and re-evaluated model predictions. To enhance comparability with other studies, we used the “survivalROC” R package (Version 1.0.3.1) to convert C-index values into AUC (32). The association between the ProSig proteins and dementia subtypes (AD) and vascular dementia (VaD)) was performed using Cox-PH models adjusted for basic risk factors (M1) as a sensitivity analysis.

#### Imaging associations

We used linear regression models to investigate the associations between the ProSig or individual ProSig proteins, and global IDPs, regional IDPs, Brain Age and Brain Age delta. We used the regularised coefficients from the previously calculated optimal number of proteins to generate ProSig for the imaging cohort. IDP values were scaled, and the regression models were adjusted for age, age^2^, sex and their interaction terms (age×sex, age^2^×sex), the MRI centre, total intracranial volume and the time difference between baseline assessment and the imaging assessment. *p*-values were adjusted for the FDR.

#### Mediation analysis

To explore whether ProSig or any individual ProSig protein mediated the effect of genetic (*APOE-ε4* allele count and an AD PRS) and modifiable risk factors (education, BMI, diabetes, hypertension, Townsend index, smoking and drinking status) on dementia risk, we conducted mediation analyses following the framework described by Baron and Kenny and additionally estimated indirect effects (33,34). The Baron and Kenney criteria investigate the presence of a mediation effect using the following steps: (1) establish an association between the predictor X (*APOE-ε4*, AD PRS or lifestyle risk factors) and the outcome Y (incidence of dementia), (2) establish an association between the predictor X and the mediator M (ProSig or ProSig protein levels), (3) establish an association between the mediator M and the outcome Y while controlling for the predictor, and (4) observe a reduction or elimination of the association of the predictor and the outcome Y when the mediator M is included in the model, indicating partial or total mediation. In the first step, we used Cox-PH regression to assess the association between each risk factor (genetic or modifiable) and time to incident dementia. We then performed linear regressions between each risk factor and ProSig or individual protein levels. In step three we performed a Cox-PH regression investigating the association between genetic and modifiable risk factors, and proteins with time in years to the incidence of dementia. Protein coefficients were recorded while adjusting for each risk factor. Proteins that were not significantly associated with incident dementia after adjustment for the risk factors (*p* >=0.05) were excluded from further analyses (step 3). As a fourth step, the coefficients for the association of the risk factors with time to incident dementia when adjusting for proteins were recorded. If the association with dementia was still significant after protein adjustment, and of the same magnitude as the association between dementia and the risk factor variables without protein adjustment, the mediation criteria were not met, and we concluded that the association of the risk factors with dementia was not mediated by these proteins. Mediation analyses were performed for each protein separately and all regression models were adjusted for age, age^2^, sex and their interaction terms (age×sex, age^2^×sex). If partial or full mediation was observed, the indirect effect of the risk factor was estimated using the product of coefficients method (path a × path b); the confidence intervals were obtained using bootstrapping with the R package “boot” (Version 1.3-28) (35). The total effect was estimated as indirect effect + direct effect (step 3, coefficient c’ for the variables).

#### Mendelian randomisation

For AD, we used summary statistics from individuals of European ancestry as described by Wightman et al (36); excluding UKB participants with proxy phenotyping based on family history due to its potential to bias effect direction in downstream analyses (37). We included AD in our MR analyses to increase power due to the large AD GWAS studies available. For ACD, we used summary statistics from the FinnGen study (38). Primary protein quantitative trait loci (pQTL) summary statistics for all signature proteins were obtained from the study by Sun et al (N = 54,219) (14). All analyses were conducted using cis-pQTLs (within 1 Mb of the gene). For several proteins, no overlapping variants, or variants in high linkage disequilibrium (LD; r² > 0.8), with the AD or ACD GWAS were identified when using cis-pQTLs from the UKB. Therefore, these proteins were instrumented using cis-pQTLs identified in an Icelandic cohort (N = 35,559) which had proteomic measurements using the SomaLogic platform (39). Analyses were restricted to the autosomes only. All variants had an F-statistics> 10, ensuring sufficient instrument strength.

MR was performed using the “TwoSampleMR” R package (Version 0.6.9) (40). For proteins that had multiple cis-pQTLs, independent instrumental variables (IVs) reaching genome-wide significance were selected as exposures and clumped using PLINK based on the 1000 Genomes Project European reference panel. Clumping was performed at an r^2^ threshold of 0.01. Prior to MR, exposure and outcome datasets were harmonised to ensure alignment of effect alleles and strand-ambiguous palindromic variants with MAF > 0.42 were excluded. Causal estimates were obtained using the Wald ratio method when only a single instrument was available. For proteins with multiple instruments, inverse-variance weighted MR with fixed effects was applied. MR analyses were conducted separately for each protein. An FDR-adjusted *p*-value < 0.05 was considered statistical significance.

## Results

### Study cohort

Out of the 502,356 participants of the prospective UKB study, 255 people had dementia at baseline (prevalent cases) and were excluded from further analyses. After quality control, we obtained a total study population of 43,685 people including 1,109 cases of incident dementia including 577 AD and 233 VaD, and plasma proteomic data for 728 inflammatory proteins (Fig. 1). Participants with incident dementia were on average older at baseline (average age of 64.5 versus 56.6 of the control). They were also more likely to be male, heterozygous or homozygous carriers of *APOE-ε4* alleles, have a higher BMI (27.8 versus 27.4 in non-dementia), have hypertension or type 2 diabetes, be less likely to have completed secondary education and had a higher deprivation index (Supplementary Table 1).

**Figure 1:**
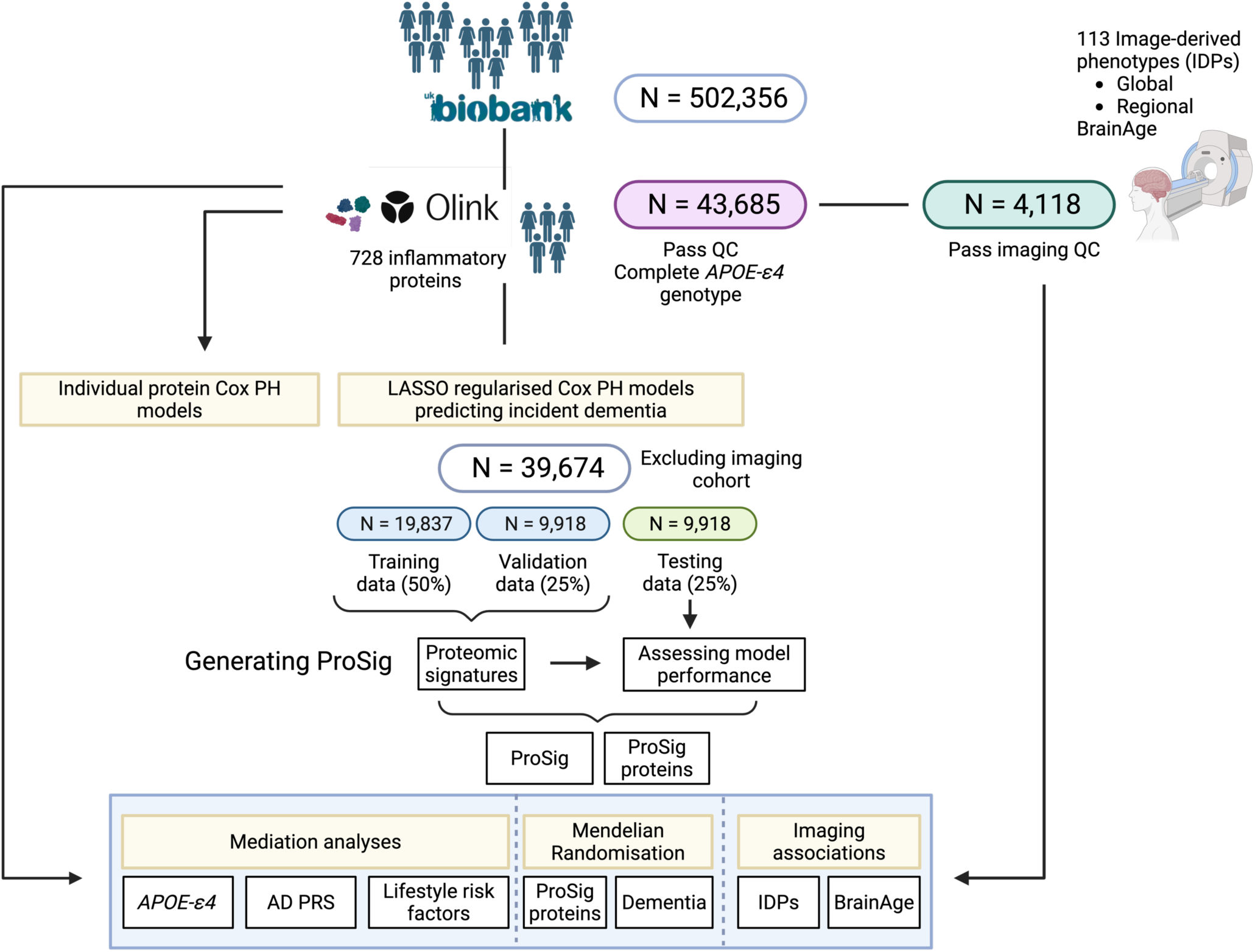
Overview of the study design and analytical pipeline. The data used in the present study is from a subset of the UK biobank population that had proteomic measurements passing our quality control (QC) and complete *APOE-ε4* genotype (N = 43,685). A further subset of these individuals have quality-controlled brain scans and are excluded from machine learning based prediction models (N = 4,118). To generate proteomic predictors of incident dementia, the study cohort (N = 39,674) was split into a training (N = 19,837), validation (N = 9,918) and testing subset (N = 9,918). LASSO regularised Cox PH models were used for feature selection and coefficient shrinkage of 728 inflammatory proteins in the training data across 100 iterations. Proteomic signatures (ProSig) were created by selecting proteins with the highest absolute mean coefficients across all iterations. The optimal number of proteins included in the ProSig was determined based on its predictive performance in the validation dataset. The final predictive performance of the ProSig was determined in the testing data. Individual level Cox proportional hazard (PH) models and mediation analyses for 728 inflammatory proteins or ProSig proteins were performed in the entire study cohort, including the imaging subset (N = 43,685). To avoid overfitting, mediation analyses for ProSig were performed only in the testing data. The association between ProSig and ProSig proteins with 113 image-derived endophenotypes (IDPs) and BrainAge was investigated in the imaging subset. Mendelian randomisation analyses were performed between protein qualitative traits of the ProSig proteins and genome wide association study summary statistics of Azheimer’s disease or all-cause dementia.

### Association of individual proteins with incident dementia

Associations between plasma inflammatory protein levels measured at baseline – up to 17.1 years prior to a dementia diagnosis – and the incidence of dementia was assessed using Cox-PH analyses (Fig 2A). In M1, adjusted for age, sex and their interactions, 218 of the 728 proteins were significantly associated with incident dementia (*p_FDR_* < 0.05; Fig. 2B, Supplementary Table 2) with HR ranging from 0.57 for apolipoprotein E (APOE) (95% CI = [0.54, 0.60], *p_FDR_* = 3.01 x 10^-8^) to 1.38 for myeloid and erythroid nuclear termination stage-specific protein (MENT) (95% CI = [1.30, 1.46], *p_FDR_* = 4.37 x 10^-23^). In M2, further adjusted for *APOE-ε4* carrier status, 196 out of 728 proteins passed the *p_FDR_* < 0.05 threshold with the HR ranging from 0.83 for C-Type Lectin Domain Family 3 Member B (CLEC3B) (95% CI = [0.78, 0.88], *p_FDR_* = 4.49 x 10^-7^) to 1.29 for plasminogen Activator, Urokinase Receptor (PLAUR) (95% CI = [1.20, 1.38], *p_FDR_* = 1.25 x 10^-10^) (Fig. 2C, Supplementary Table 2). 170 proteins remained significant after *APOE-ε4* allele adjustment; however it also attenuated the significance of 48 proteins. We also observed that 26 proteins, which showed no association with incident dementia in M1, became significantly associated with incident dementia after adjusting for *APOE-ε4* alleles. In M3, (M1 with additional adjustment for ethnicity and modifiable risk factors), we identified 198 associations with HR ranging from 0.59 (APOE, 95% CI = [0.55, 0.62], *p_FDR_* = 6.98 x 10^-72^) to 1.45 (MENT, 95% CI = [1.36, 1.54], *p_FDR_* = 3.23 x 10^-30^). M4 was adjusted for all covariates, including *APOE-ε4* genotype and showed 138 significant associations, with HRs varying from 0.84 (95% CI = [0.80, 0.90], *p_FDR_* = 1.47 x 10^-4^) for Lecithin–cholesterol acyltransferase (LCAT) to 1.21 (PLAUR, 95% CI = [1.13, 1.31], *p_FDR_* = 1.27 x 10^-4^) (Fig. 2E; Supplementary Table 2). Overall, the most significantly associated proteins between M1 and M3, as well as between M2 and M4, were the same. The two proteins displaying the highest overall associations across all models were PLAUR and Cluster of Differentiation 276 (CD276). We examined the proportionality of hazard assumptions using Schoenfeld residuals, which revealed constant HR for most proteins (Supplementary Table 2).

**Fig. 2.**
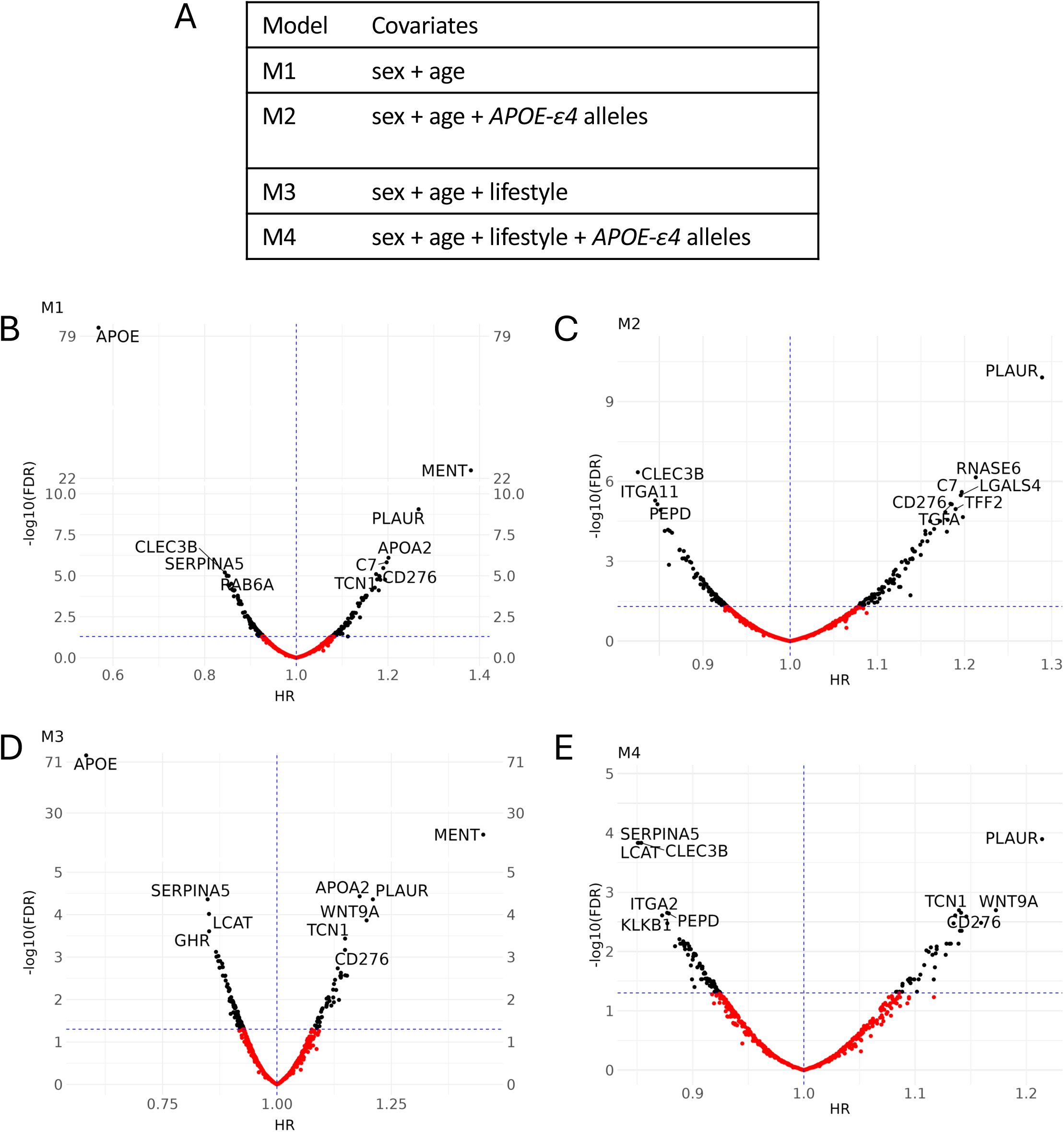
Individual inflammatory protein association with incident dementia. **(A)** The table summarises the covariates, besides proteins levels, that were taken into adjustment in each Cox PH model. (B-E) Volcano plots demonstrating HR (x-axis) against corrected *P* values (y-axis) in models adjusted for sex and age (M1; B), sex, age and *APOE-ε4* alleles (M2; C), sex, age and lifestyle risk factors (M3; D) and sex, age, lifestyle risk factors and *APOE-ε4* alleles (M4; E). The lifestyle risk factors include BMI, ethnicity, education, deprivation, hypertension, diabetes, alcohol consumption and smoking status. The proteomic associations with dementia onset are significant in proteins indicated in black (*p_FDR_* <0.05) and non-significant in those indicated with red (*p_FDR_* ≥ 0.05). The horizontal lines indicate *p_FDR_* = 0.05 and the vertical lines show a hazard ratio of 1. The proteins with the lowest *p_FDR_* have been labelled.

### Inflammatory proteomic signature of dementia

Using LASSO-penalised Cox-PH models in the training set, we identified 80 proteins with non-zero regularised coefficients at least in one iteration. We constructed ProSig by sequentially adding proteins in groups of five, ordered by absolute coefficients, and found that the C-index plateaued after the first 20 proteins (C-index = 0.81; Fig. 3A, Supplementary Table 4). The addition of the ProSig significantly increased the predictive power of Cox-PH models that were adjusted for sex, age, ethnicity and modifiable risk factors (M1 and M3; Fig. 3B-3C), as well as the C-index of the model adjusted for sex, age and *APOE-ε4* alleles (M2). Finally, the inclusion of ProSig in the fully adjusted model (M4) resulted in an increase in the C-index, though the improvement was not statistically significant (Fig. 3C). Model performance remained largely consistent in sensitivity analyses restricted to dementia cases with onset within 5, 10, and 15 years (Supplementary Table 5). Individual-level regression analyses (M1) showed that 12, 11 and 6 ProSig proteins were associated ACD, AD and VaD respectively (Supplementary figure 1).

**Fig. 3.**
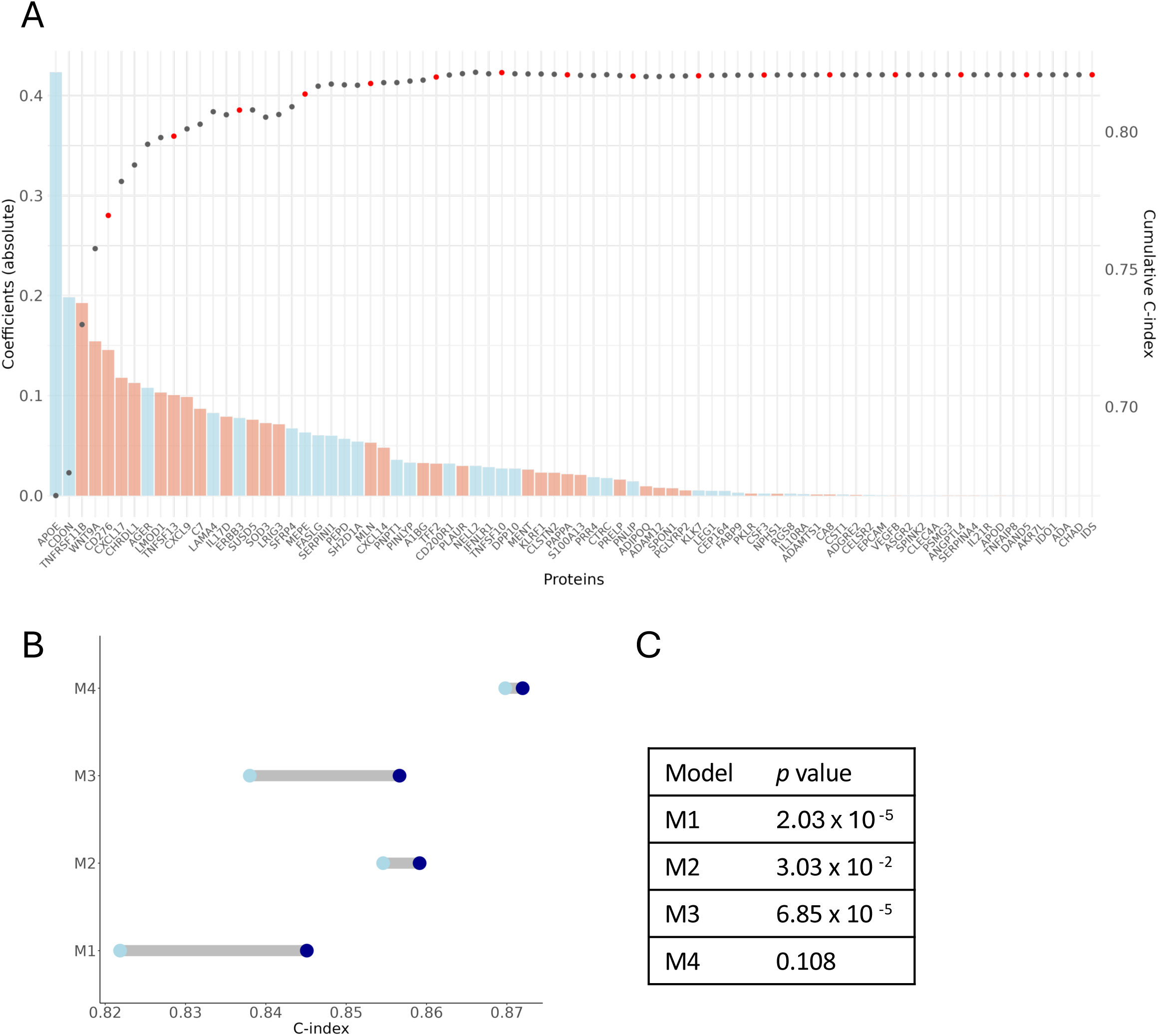
Optimisation of proteins included in the generation of the proteomic signature (ProSig). **(A)** The dot plot illustrates the C-index values (y-axis) of Cox proportional hazards models adjusted for the ProSig in the validation sample, with proteins incrementally added to the ProSig based on their highest absolute coefficients. Red dots highlight every fifth increment. The bar plot illustrate the mean absolute LASSO-regularized coefficients (y-axis) of ProSig proteins (x-axis) across 100 iterations, with blue bars representing negative coefficients and red bars representing positive coefficients. **(B)** Changes in C-index values across Cox PH models with varying adjustments, following the inclusion of the 20-protein ProSig. M1 is adjusted for sex and age. M2 is additionally adjusted for *APOE-ε4* alleles. M3 is adjusted for age, sex and lifestyle risk factors (BMI, ethnicity, education, deprivation, hypertension, diabetes, alcohol consumption and smoking status) and M4 is adjusted for sex, age, lifestyle risk factors and *APOE-ε4* alleles. The light blue circles show the C-index without ProSig and the dark blue circles show the C-index with ProSig. **(C)** Statistical significance of differences in C-index values observed after adjusting for the ProSig in Cox PH models.

As a sensitivity analysis, we stratified proteins in the ProSig based on the dementia subtype and investigated individual protein associations with AD and VaD (Supplementary figure 1). There was a higher overlap between ProSig proteins individually associated with AD and ACD (n = 10) versus VaD and ACD (n = 5). CHRDL1 was uniquely associated with incident VaD whilst IL17D was only associated with incident AD.

### Association between ProSig, key metabolites and brain image-derived phenotypes

We observed limited associations between the ProSig or individual proteins within ProSig with global IDPs, with CXCL17 showing the most widespread associations including reduced gray matter, cerebral white matter and cortical volumes (Fig. 4A, Supplementary Table 6). The ProSig was associated with the volume of the cerebrospinal fluid (CSF; β = -0.015, *p_FDR_* = 6.71 x 10^-3^, *p_FDR_*). In the regional brain volume analysis, the majority of significant associations (*p_FDR_* < 0.05) were observed with subcortical structures. The key regions with significant reductions in volume were the ventral diencephalon which encompasses several structures including the thalamus and hypothalamus, the thalamus proper, the hippocampus, the cerebellum white matter and the cerebellum cortex. The proteins with highest association with these regions were CXCL17, tumour necrosis factor receptor superfamily member 11B (TNFRSF11B) and cell adhesion associated, oncogene regulated (CDON) (Fig. 4B).

**Fig 4.**
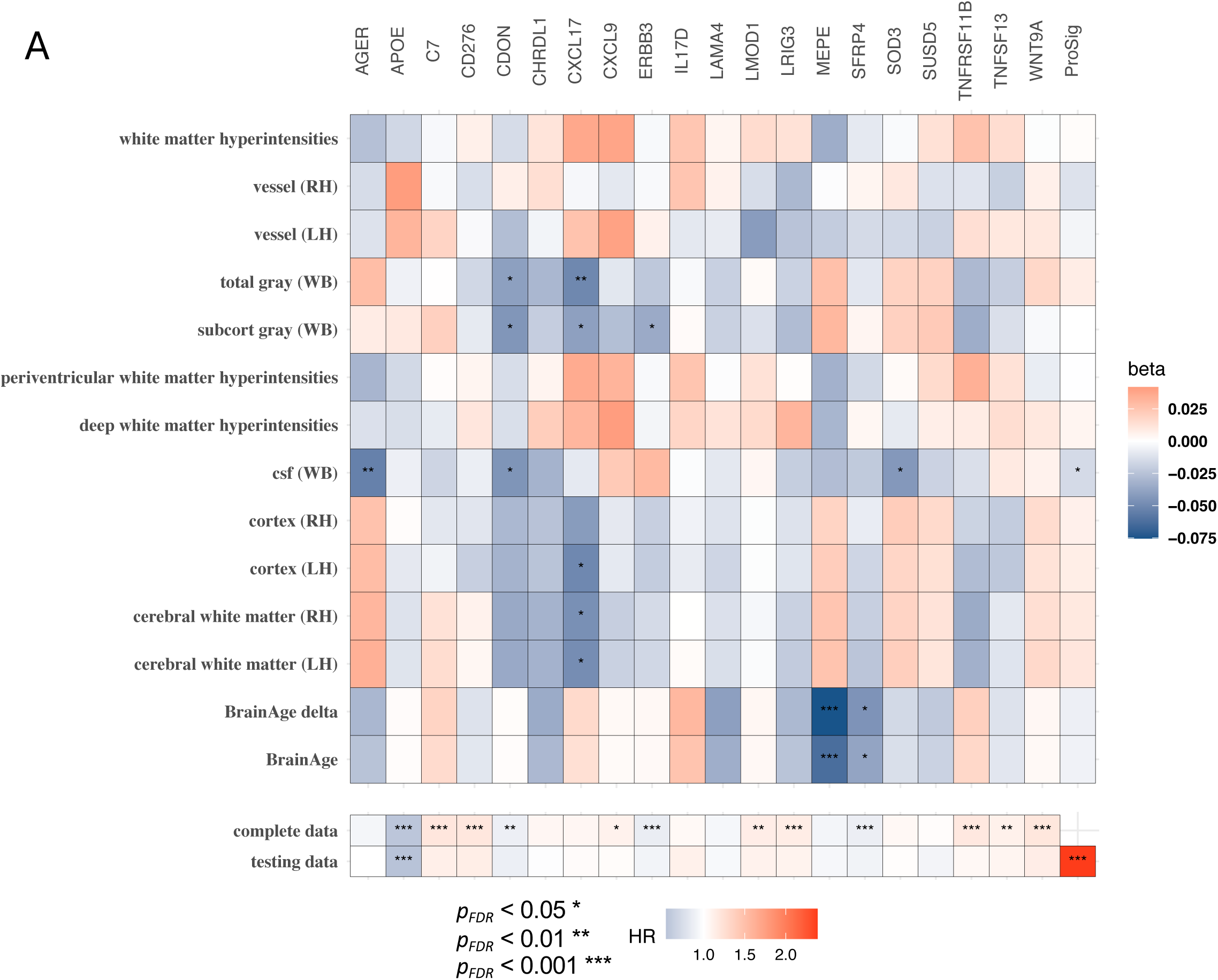
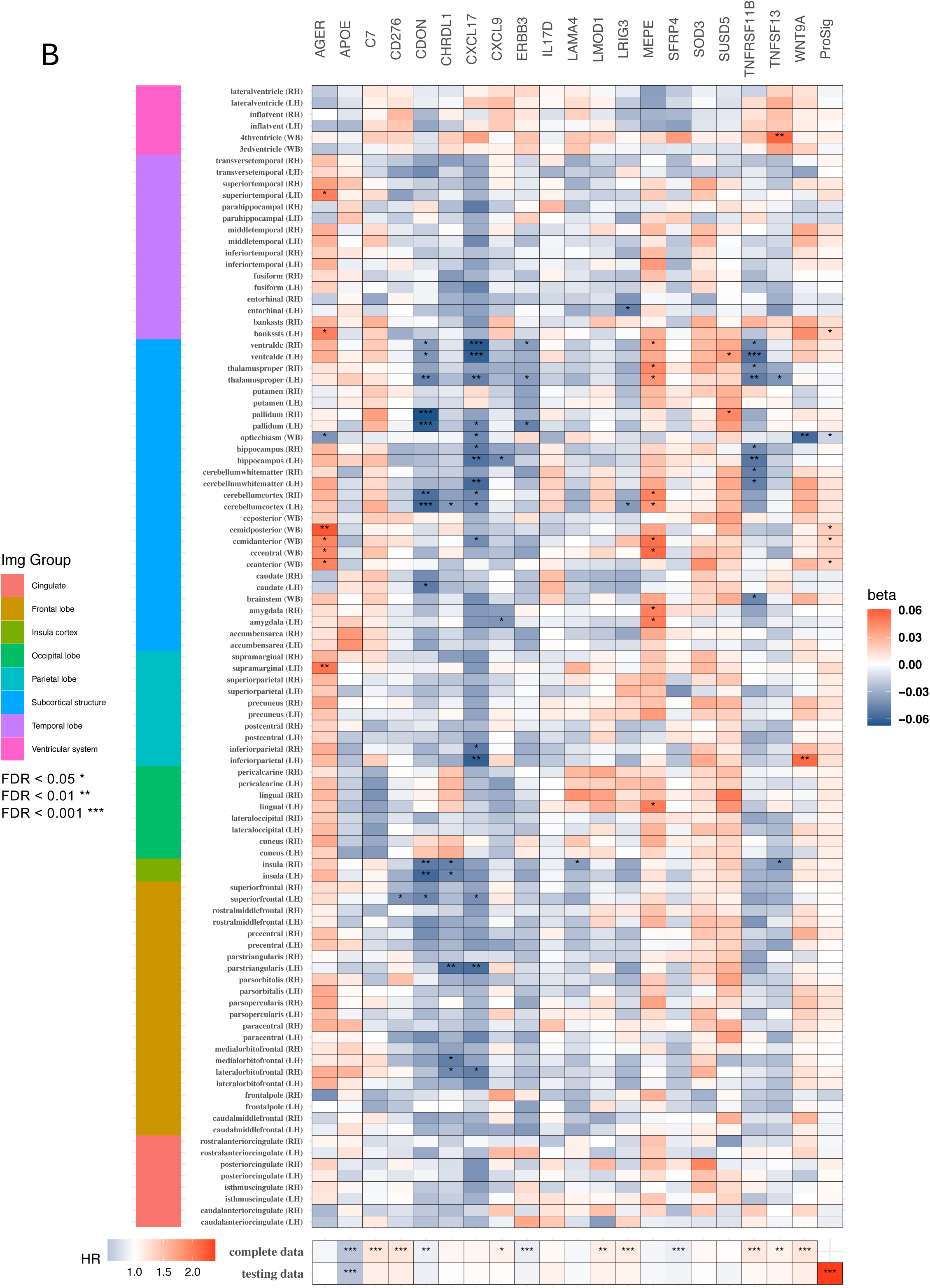
The association between proteomic signature (ProSig) and ProSig proteins with MRI-derived brain volumes. **(A-B)** The top panels show the associations between global (A) and regional (B) image-derived phenotypes (y-axis) and ProSig proteins or ProSig (x-axis). Bottom panels show the Cox PH-derived association between ProSig proteins or ProSig with incident dementia in the test data and all data. WB: whole brain; RH: right hemisphere; LH: left hemisphere.

### Mediation analysis

We performed mediation analysis using the 20 ProSig proteins in the whole study population (Supplementary Table 7). Το avoid overfitting, we investigated the mediating effect of the ProSig in the test data (Supplementary Table 8). Of the 180 associations examined (9 predictors and 20 proteins), 71 instances showed significant protein-mediated effects for 8 predictors. Most of these effects were for diabetes (n = 14) and hypertension (n = 13). Although most were marginal, 21 mediatory effects accounted for 5% or more of the total effect. Among these, TNFRSF11B emerged as a key mediator, influencing over 5% of the effects of smoking status (11.45%), Townsend index (9.41%), diabetes (7.48%), education (7.31%) and hypertension (6.64%) on dementia (Fig. 5A). Overall, APOE exhibited the strongest mediatory effect, accounting for 22.38% of the impact of diabetes on dementia (Fig 5B). Consistent with this, the ProSig mediated 41.35% of the effect of diabetes. However, this mediation was predominantly driven by the APOE protein, which carries the highest weight in the score. When APOE was excluded from the ProSig, the mediation proportion of 41.35% dropped to 15.33%. Moreover, complement component 7 (C7) mediated 11.9% of the effect of smoking and 9.7% of the effect of Townsend index on dementia (Fig 5. A-B). Finally, we found five proteins (ERBB3, sFRP4, TNFSF13, CXCL9 and TNFRSF11B) that significantly mediated the effect of the *APOE-*ε4 genotype on dementia, and one protein (AGER) that partially mediated the effect of an AD PRS; however, all mediatory effects were marginal (less than 1%).

**Fig 5.**
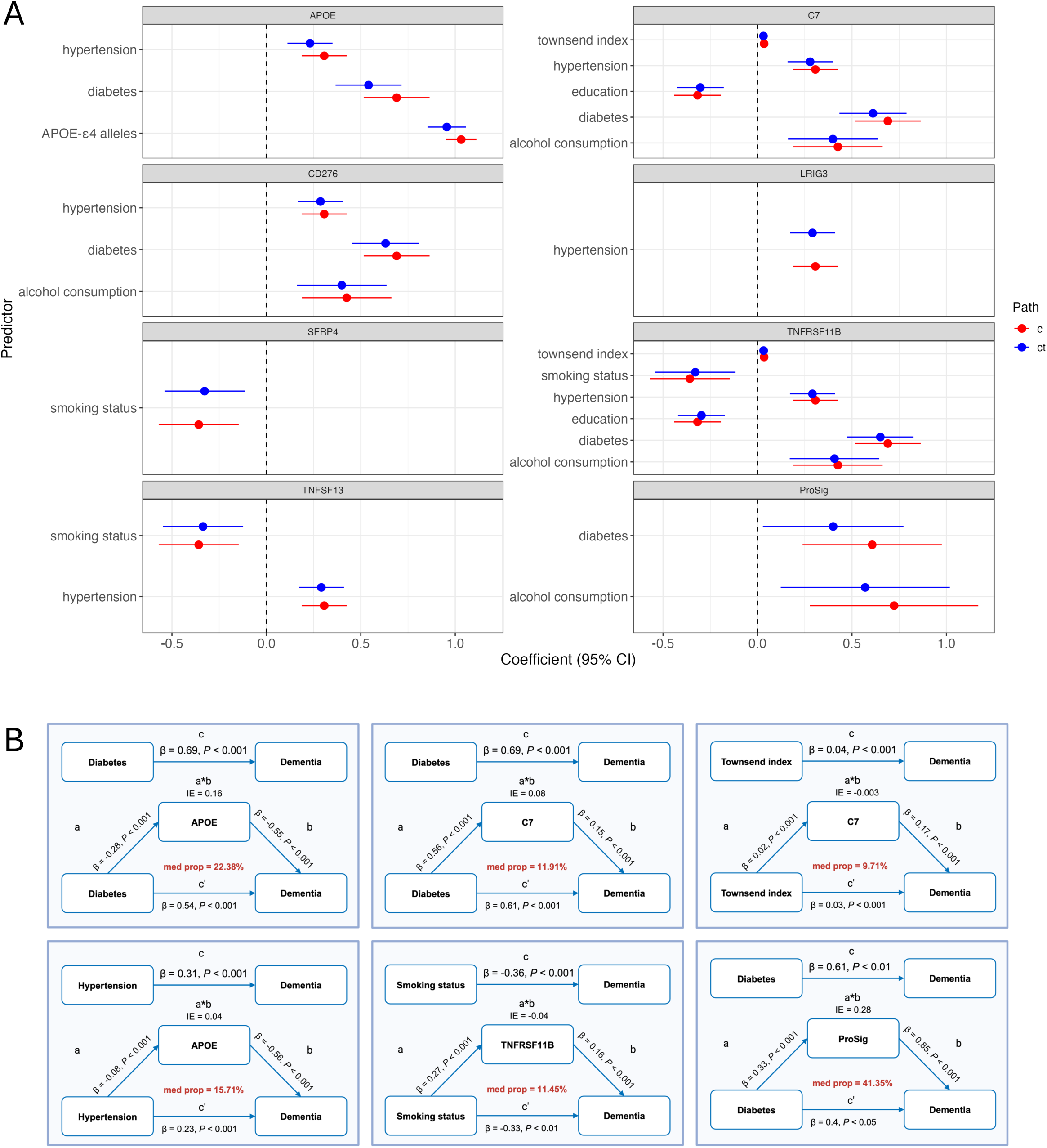
Mediating effect of the ProSig proteins and the ProSig on the relationship between genetic and lifestyle risk factors with dementia onset. **(A)** Forest plots showing the direct (c’) and total effect (c) in the survival association between the ProSig proteins or ProSig and dementia predictors. The mediation proportion of 5% or more has been shown in this figure. **(B)** Schematics of a, b, c and c’ paths in mediation analyses with the regression coefficients at each step. The plots show mediation proportions that are at 10% or more. IE: indirect effect; med prop: mediation proportion. **(A, B)** The association with ProSig is only examined in the test data and the remaining associations are explored in the complete data.

### Mendelian Randomisation

In two-sample MR analyses, we identified nominally significant inverse associations for tumour necrosis factor ligand superfamily member 13 (TNFSF13) with both AD (β = -0.094, se = 0.04, *p* = 0.012) and ACD (β = -0.104, se = 0.04, *p* = 0.013), and for interleukin (IL)-17D with ACD (β = -0.071, se = 0.03, *p* = 0.032; (Supplementary table 9) . However, these results should be interpreted cautiously as they did not pass FDR correction and were only nominally significant.

## Discussion

Here, we applied a triangulation framework—integrating observational analyses, machine learning, mediation analyses and MR—to comprehensively evaluate the contribution of systemic inflammation to dementia risk. We first developed a plasma inflammatory proteomics signature comprising of 20 proteins – ProSig – that predicted incident dementia beyond traditional risk factors. We identified associations with subcortical volumes, including the thalamus and hippocampus, key brain regions critically implicated in dementia (41), as well as a lower Brain Age delta. Mediation analyses further showed that ProSig and some of the individual ProSig proteins mediated the effects of genetic and modifiable risk factors on dementia risk. Finally, MR analyses provided evidence for a protective causal association of two ProSig proteins with both ACD and AD.

In univariable Cox-PH analyses, we found that 30% of the inflammatory proteins were associated with incident dementia (*p_FDR_* < 0.05). Previous research in the same cohort using cardiometabolic, inflammation, neurology and oncology Olink panels (1,472 total proteins including the 368 inflammatory proteins used here) had shown approximately 4% of the investigated proteins were associated with incident VaD and around 1% significantly associated with AD (16). To enhance comparability, when we applied a similarly calculated threshold, we found that 7% of the inflammatory proteins were associated with incident dementia.

For our prediction models, we used machine learning with independent data splits to identify a protein signature for incident dementia, aiming to minimise both overfitting and the number of proteins, while preserving predictive performance. Our results, generated from only inflammatory proteins, are comparable to and align with previous research and further demonstrating that incorporating a proteomic signature can improve predictive accuracy beyond risk factors and clinical markers (12,16). For example, earlier studies have reported substantial increases in the AUC (5–6%) when protein scores were integrated into Cox-PH models, including both minimally adjusted models and those accounting for more complex risk factors (AUC ∼ 0.90 or C-index of 0.89) (12,16). Similarly, we saw an improvement in the AUC with the addition of the ProSig to models adjusted for sex, age and modifiable risk factors (AUC 0.86 vs 0.89 or C-index 0.84 vs 0.86, *p* = 6.51 x 10^-5^) without using clinical factors (Supplementary Table 5). However, we observed a modest increase in the prediction performance of the fully-adjusted model with the addition of the ProSig which was not statistically significant (AUC 0.89 vs 0.90 or C-index 0.870 vs 0.872, *p* = 1.08 x 10^-1^), highlighting that the proteomic signature does not offer a substantial improvement in prediction compared to the *APOE-ε4* genotype and modifiable risk factors together – something that was not explored in previous studies (12,16,42,43). This finding suggests that proteins in the ProSig may partly capture the same biological effects as *APOE-ε4*. Recently, a study from the Global Neurodegeneration Proteomics Consortium (GNPC) further emphasised the importance of investigating inflammatory proteins in neurodegenerative disease, showing that *APOE-ε4* carriers display proteomic signatures enriched in pro-inflammatory markers (44). Our mediation analyses suggested a minuscule mediating role of ProSig proteins in the *APOE-ε4*–dementia relationship, and the lack of predictive improvement from adding *APOE-ε4* may reflect potential confounding. Moreover, as the UKB (Olink) and the GPNG (Somalogic) use different proteomics platforms with some overlap (45), future work should focus on systematically investigating the interplay of *APOE-ε4* and inflammatory proteins on dementia risk across different cohorts and platforms.

From the 20 proteins identified in our signature, we found that APOE, AGER, C7, CDON, and CXCL9 were previously identified in dementia signatures in large-scale observational studies and their roles in neurodegeneration is relatively well explored (16,46–50). Another study from the GNPC used LASSO regularisation to identify a 256-protein global signature of dementia severity based on the Clinical Dementia Rating global scores across individuals with AD, Parkinson’s disease and frontotemporal dementia (45). Two proteins from their signature (C7 and sFRP4) overlapped with our findings. Six proteins (CD276, sFRP4, TNFRSF11B, ERBB3, CHRDL1, and CXCL17) have been reported in dementia signatures from larger studies, though their mechanistic roles remain unclear (16,43,45,51).Four proteins (TNFSF13, LAMA4, and WNT9A ) have been previously associated with dementia in smaller cohorts, with some mechanistic evidence (52–54), whereas SUSD5 was also observed in smaller studies but without proposed mechanisms. (52,55). Finally, SOD3 and LMOD1 were identified in dementia animal models or postmortem studies but have not been replicated in case-control or cohort studies (56,57).

To detect subtle brain changes before dementia symptoms emerge and to link molecular pathways to brain pathology, we investigated associations between ProSig proteins and IDPs. We found only few associations between the ProSig and IDPs, likely due to heterogeneous protein effects neutralising each other in the composite. Similarly, individual proteins showed limited associations with global brain volumes, possibly reflecting region-specific effects diluted at a larger scale. To gain deeper insights into specific structural changes, we examined regional IDPs. We mostly found associations with gray matter and subcortical regions, including the hippocampus, ventral diencephalon, thalamus proper, cerebellar white matter, and cerebellar cortex, bilaterally. These structural changes are implicated in dementia and suggest a potential role for these proteins in early neurodegenerative processes (58–62). Moreover, these associations best align with AD pathology, compared to fewer significant associations with white matter, that reflects vascular changes.

Overall, we found different patterns in the association of the ProSig proteins with brain phenotypes and dementia. For example, whilst TNFRSF11B and CDON were both associated with reduced subcortical brain volumes, only TNFRSF11B was associated with a significant higher risk for dementia, supporting the involvement of the protein in pathways that accelerate cognitive decline (51,52); whereas CDON was protective against dementia, which could suggest a protective role or compensatory mechanism despite the observed structural changes. This could be supported by the role of CDON in neurogenesis, oligodendrocyte differentiation and myelination (63). On the other hand, CXCL17 was associated with reduced subcortical volumes but not with incident dementia. This may reflect involvement in early disease processes, reduced neural reserve, or age-related changes unrelated to dementia. Interestingly, CXCL17 has previously been studied in the context of proteomic aging (64). Additionally, proteins such as CD276 showed strong associations with incident dementia across sequentially adjusted models but were not linked to structural brain changes. We also found two proteins, MEPE and sFRP4, associated with a lower Brain Age delta, with sFRP4 being also protective against dementia. sFRP4 is an antagonist of the Wnt signaling pathway, a prominent pathway for synaptic maintenance and plasticity in the adult brain, the downregulation of which has been linked to AD and neuronal instability (65,66). While other sFRP members have been implicated in dementia, evidence for sFRP4 is lacking (66). Notably, sFRP show biphasic effects – acting as inhibitors or potentiators of Wnt signalling depending on their concentration and cellular context – and studies on related family members in neurogenesis have produced mixed findings (66). Here we found that sFRP4 may have neuroprotective properties that help delay neurodegeneration and brain aging, increase resilience and prevent dementia.

Next, we examined whether ProSig proteins mediated the effects of genetic and modifiable risk factors on dementia, aiming to better understand their potential role in underlying disease pathways and biological mechanisms. We observed that some ProSig proteins showed statistically significant mediation of genetic risk (both *APOE-ε4* and AD PRS), but the effect sizes were negligible (less than 1%). Amongst all proteins and modifiable risk factors, the highest mediation proportions were observed for APOE (22.48%) and C7 (11.91%) with diabetes, APOE with hypertension (15.71%) and TNFRSF11B with smoking status (11.45%). Finally, we investigated the mediatory effects through the ProSig in the test data only and we observed that the ProSig mediated 41% of the effect of diabetes on dementia. However, these associations were likely inflated due to the smaller sample size of the test sample, and were mainly driven by APOE, given the observed individual mediatory effect with diabetes and the high influence of the APOE protein on the ProSig. Some of the mediatory effects of individual proteins are biologically plausible, warranting further investigation, such as the mediating effect of TNFRSF11B on the association between smoking and dementia risk. Smoking leads to impairment in the vascular endothelial function and induces an increase in pulmonary arterial blood pressure (67). TNFRSF11B is upregulated in vascular cells in pulmonary arterial hypertension ((PAH) – a rare condition caused by endothelial dysfunction in the pulmonary arteries, preventing apoptosis and promoting proliferation of these cells (68). Nevertheless, observational mediation—even in our study, where protein samples were collected years before dementia onset—can be limited by potential unmeasured confounding, temporal ambiguity, reverse causation, measurement error, collider bias, difficulty handling multiple mediators, and restricted causal interpretability (56). Further research is therefore needed to elucidate the causal nature of these relationships.

Using MR, a method that can overcome some of the limitations of observational epidemiology, we finally showed that two proteins TNFSF13 and IL17D may have potentially causal associations with AD and ACD. The potential protective causal effect for TNFSF13 on AD and ACD suggests neuroprotective role of the protein under genetically driven and life-long expression. However, in our observational survival models, higher circulating levels of TNFSF13 were associated with an increased risk of dementia in models adjusted for sex, age and the *APOE-ε4* genotype (HR = 1.13, CI = [1.06, 1.20], *p* = 0.0036 in M2). The association was attenuated when models were additionally adjusted for modifiable risk factors (HR = 1.08, CI = [1.01, 1.15], p = 0.070 in M4); suggesting observational associations may be confounded by these variables. However, this apparent discrepancy should be interpreted cautiously; the MR effect is nominal and MR reflects a lifelong, genetically driven protective effect of higher TNFSF13 levels against dementia and AD, whereas the observational models capture cross-sectional circulating levels that may be elevated in later life due to environmental exposures or disease-related immune activation. Previous observational studies have reported an elevation in TNFSF13 in CSF of those with AD (69). Moreover, MR analyses in other studies have shown causal protective associations between TNFSF13 gene expression and AD (70). Similarly, we did not find any significant associations between IL17D and incident dementia or neuroimaging phenotypes. This may suggest that the protective effect of IL17D has not been captured in the single time point protein measurement, or competing processes whereby the protective effect of IL17D is attenuated. Blood IL17D was part of a proteomic panel that accurately differentiated individuals with dementia from healthy controls (71), associated with increased risk of dementia and reduced cognitive performance (71). Furthermore, IL17D is upregulated in the astrocytes of AD patients and might be part of the brain’s attempt to cope with disease pathology (72). Taken together, findings suggest a potential protective role for IL17D against dementia that warrants further investigation.

One of the key findings of our study was the role of TNFRSF11B, also known as OPG, in early brain changes, its ability to predict dementia, and its partial mediation of the effect of smoking on dementia risk. TNFRSF11B is expressed in microglial cells and is involved in regulating inflammatory responses in the central nervous system, as well as contributing to vascular endothelial dysfunction and degeneration of the blood-brain barrier (73,74). TNFRSF11B has previously been implicated in dementia in different cohorts (16,75,76). In cognitively healthy individuals from the Framingham Offspring cohort, higher OPG levels were associated with smaller total cerebral brain volume (77). Murine studies have suggested TNFRSF11B is an early pathological marker of AD (78). Notably, TNFRSF11B has already been successfully targeted with a human antibody for vascular remodelling in rodent models for pulmonary arterial hypertension (PAH), making the protein a promising candidate for future dementia intervention (79). Our findings provide robust population-level evidence supporting a role for TNFRSF11B in early dementia pathology and preclinical structural changes in key dementia-related brain regions-as well as a potential mediatory role in disease pathways. Nonetheless, we did not find evidence for a causal association between TNFRSF11B and dementia.

Our study has some limitations. Dementia is an umbrella term encompassing a variety of diagnoses, but distinguishing between dementia subtype remains challenging due to lack of objective tests and reliance on clinical judgement (80). Particularly, dementia subtyping in UKB is thought to be imprecise, especially for VaD, and ACD cases may be significantly under-ascertained when relying solely on hospital or death records (17). To address this, and to maximise statistical power, we calculated proteomic signatures for all-cause dementia and looked for AD-specific associations in our MR analyses. Additionally, our data does not provide details on the tissue of origin of plasma proteins nor their activity, which could be addressed in future studies by integrating tissue-specific datasets or validation in the CSF. We have based our definition of inflammatory proteins on the Olink panels from the UKB data; however, this definition is not unanimous across all data banks. The proteins have been measured only at baseline lacking follow-up measurements and although Cox-PH models account for follow-up time, using only baseline protein measurements may overlook temporal changes that could influence risk prediction; similarly, brain images were available at a single time point. We found a number of proteins strongly associated with dementia in univariable Cox-PH models (e.g. PLAUR and MENT) that were not part of our signature, which warrant further investigation in future studies. Although we used available data from all participants, adjusting for ethnicity, the UKB proteomic data lacks diversity and is predominantly from participants of white backgrounds and well educated who are healthier than the general population (81). Furthermore, the proteomics and imaging sub-samples in the UKB were not randomly selected; those who attended follow-up appointments were more engaged with the study than the full cohort and were usually healthier and had lower deprivation. Notably, the imaging analyses includes the healthiest participants, and their proteomic data were not used to generate the ProSig. Mediation analyses were limited by the smaller test sample size and the strong influence of APOE, which may have inflated the observed mediatory effects. The data used for our MR analyses were also from individuals of European ancestry. Due to the missing cis-pQTL data for some proteins in the UKB dataset, we used instruments derived from an Icelandic cohort measured with the SomaLogic platform. This introduces potential sources of heterogeneity due to differences in ancestry and proteomic measurement technologies, which may impact the comparability and interpretation of MR findings across proteins. Moreover, MR analyses require satisfaction of key assumptions, including availability of strong genetic instruments and absence of pleiotropy. While we addressed these by selecting strong instruments (F-statistics > 10), we acknowledge the results should still be interpreted with caution and warrant further triangulation. Finally, MR sensitivity analyses were limited due to the small number of instruments, making it difficult to assess potential pleiotropy—a common challenge when using pQTL data.

Future research should focus on further elucidating the causal pathways between dementia and the predictive inflammatory proteins. Additionally, identifying specific downstream pathways disrupted by the signature proteins would provide valuable insights. Finally, our findings should be validated in an independent cohort to confirm their robustness. With the emergence of new proteomic cohorts, for example GNPC, we will be able to replicate some of this work. However, only 10 of our signature proteins were measured in this consortium as the proteins were measured with the SomaLogic platform.

## Conclusion

Our findings underscore the pivotal role of inflammation in dementia pathogenesis and provide novel insights into early inflammatory mechanisms underlying neurodegeneration. Using a systematic workflow, robust methodology to reduce overfitting and triangulating evidence across different methods we have shown that the top proteins that were used to calculate our proteomic signatures capture heterogenous associations with AD and ACD which also emphasises the complexity of immune processes in dementia development. We demonstrate that alterations in subcortical brain volumes are associated with key inflammatory proteins, in some cases independent of dementia diagnosis. Moreover, several of these proteins mediate the effects of established genetic and modifiable risk factors on dementia. Of note is TNFRSF11B – a potentially important risk factor protein associated with increased risk of dementia and early brain changes.

These results highlight the potential of inflammatory biomarkers for early detection and risk stratification and may inform precision approaches to prevention and intervention, including improved targeting and recruitment for clinical trials.

## Data Availability

All data produced in the present study are available upon reasonable request to the authors

